# Measure what matters: counts of hospitalized patients are a better metric for health system capacity planning for a reopening

**DOI:** 10.1101/2020.04.19.20072017

**Authors:** Sehj Kashyap, Saurabh Gombar, Steve Yadlowsky, Alison Callahan, Jason Fries, Benjamin A Pinsky, Nigam H Shah

## Abstract

Responding to the COVID-19 pandemic requires accurate forecasting of health system capacity requirements using readily available inputs. We examined whether testing and hospitalization data could help quantify the anticipated burden on the health system given shelter-in-place (SIP) order.

We find a marked slowdown in the hospitalization rate within ten days of SIP even as cases continued to rise. We also find a shift towards younger patients in the age distribution of those testing positive for COVID-19 over the four weeks of SIP. The impact of this shift is a divergence between increasing positive case confirmations and slowing new hospitalizations, both of which affects the demand on health systems.

Without using local hospitalization rates and the age distribution of positive patients, current models are likely to overestimate the resource burden of COVID-19. It is imperative that health systems start using these data to quantify effects of SIP and aid reopening planning.

## Introduction

In order to prepare for COVID-19, health system leaders and policy-makers need to forecast future healthcare needs. A number of forecasting models have been developed and widely shared to help health care facilities and governments predict upcoming patient surges and plan accordingly ^1,2^. These models take in a myriad of inputs including population demographics, currently admitted patients, case doubling times, and the rate at which positive cases turn into hospitalizations, among other inputs ^1,3,4,5^. While there are enough models, guidance and data to provide accurate inputs to these models remain lacking ^6–8^. Currently, 95% of Americans have been asked to stay at home. Schools, businesses and community gathering activities have been closed or curtailed and plans of reopening hinge on demonstrated sustained reduction in cases, availability of testing, and enough health system capacity to treat patients requiring hospitalization without resorting to crisis standards of care ^9,10^. The need to accurately plan health system capacity is one of the six indicators cited to guide the state of California’s decision making around reopening the state’s economy ^11^. An important first step is taking stock of the currently available hospitalization and testing data and how it has evolved as the pandemic progresses.

Stanford Health Care (SHC), a large academic healthcare system, serves patients in the San Francisco Bay Area. The major hospitals that comprise SHC are located in Santa Clara County, which began mandated shelter-in-place (SIP) on midnight March 16, 26 days before we conducted the analysis presented here. SHC had extensive testing capacity starting on March 4^th^, and since then, tested anyone who had influenza-like infection symptoms, known COVID-19 positive contact, healthcare workers with a known exposure, or by physician discretion. As of April 11th, our laboratory has tested nearly 15,800 cases, and tracked hospitalization data of the test-confirmed COVID-19 cases.

Given our dual access to testing and hospitalization data, we examined whether we could reliably quantify the effects of state-mandated SIP using testing and hospitalization rates. This allowed us to quantify the divergence between the rates of positive case confirmations and hospitalizations. In addition, we identified a shift in the age distribution of new COVID-19 positive cases over the duration of the study.

## Methods

16,103 SARS-CoV-2 RT-PCR tests were performed on 15,807 patients at Stanford facilities between March 2 and April 11, 2020. Of these, 8,309 tests were performed on 7,929 patients in facilities where the patient would have been admitted to our hospital if necessary. We analyzed the fraction of tested patients that were confirmed positive for COVID-19, the fraction of those needing hospitalization, and the fraction requiring ICU admission over the 40 days between March 2nd and April 11th 2020.

Data were obtained from a combination of two sources: a daily-refreshed snapshot of the health system’s Enterprise Data Warehouse (EDW) and a twice-daily refreshed extract of all RT-PCR laboratory tests for SARS-CoV-2 at Stanford. Access to a view of the EDW was set up last year as part of Stanford Medicine’s Program for Artificial Intelligence in Healthcare^12^. The EDW view consisted of 33 tables, containing demographics, diagnoses, procedures, labs, and orders.

These tables were refreshed nightly with updated one-year historical and current hospitalization information of patients admitted in the census of the hospital. The access to the laboratory testing was provided as part of Stanford’s data science response to COVID-19^13^. The laboratory testing data consisted of details about the specimen collected, the result, the procedure of specimen collection, identifiers to link with EDW tables and symptoms documented on admission. The project to track SARS-CoV-2 test positivity and hospitalization trends was initiated as a quality assurance project aimed at enabling hospital capacity planning and IRB approval (protocol # 55544) was obtained prior to this submission summarizing the learning from this effort.

## Results

### 3.77% of COVID-19 positive patients required an ICU admission

As shown in Figure 1, out of these 7,929 tested patients, 451 (5.68%) tested positive for COVID-19 and out of these 451 cases, 59 (13.08%) were hospitalized following their test. Among the 59 hospitalized cases, 17 (28.8% of hospitalized and 3.77% of all positive cases) required ICU care. Our observed case hospitalization rate is more in line with the 12% reported nationally than the 25.5% reported in New York City (as of April 7)^14,15^. The higher case hospitalization rate in New York may be due to the fact that low testing capacity to case numbers might have led to severe cases being prioritized for testing. Our ICU rate among confirmed cases similarly matches American national reports of 2.9%, compared to the 5% reported in China or 12% in Lombardy^16,17^.

**Figure 1.**
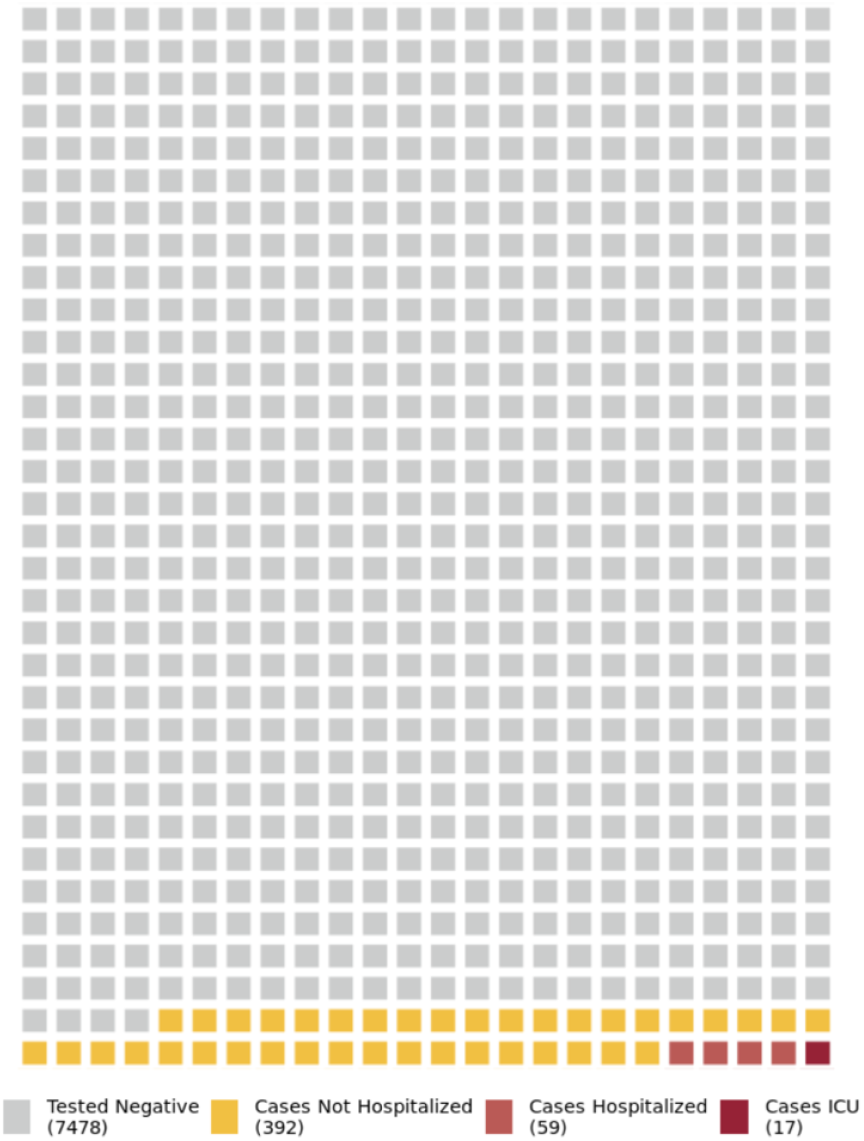
Test result as well as hospitalization outcomes of patients tested for SARS-CoV-2. Each box represents 10 patients.

### Hospitalization rate slowed within 10 days of shelter-in-place even as confirmed cases continue to rise

Over the past five weeks, we continue to see new COVID-19 cases, hospitalizations, and ICU admissions, however their rates have slowed (Figure 2). The doubling time for each metric increased from under 5 days on March 16th to over 25 days as of April 11. The slowdown in hospital admissions began within 10 days of SIP, and is more dramatic than the slowdown of confirmed cases. Forecasting models should incorporate the divergence between the rate of new COVID-19 cases versus new hospitalizations to better forecast near-term demand on the health system.

**Figure 2.**
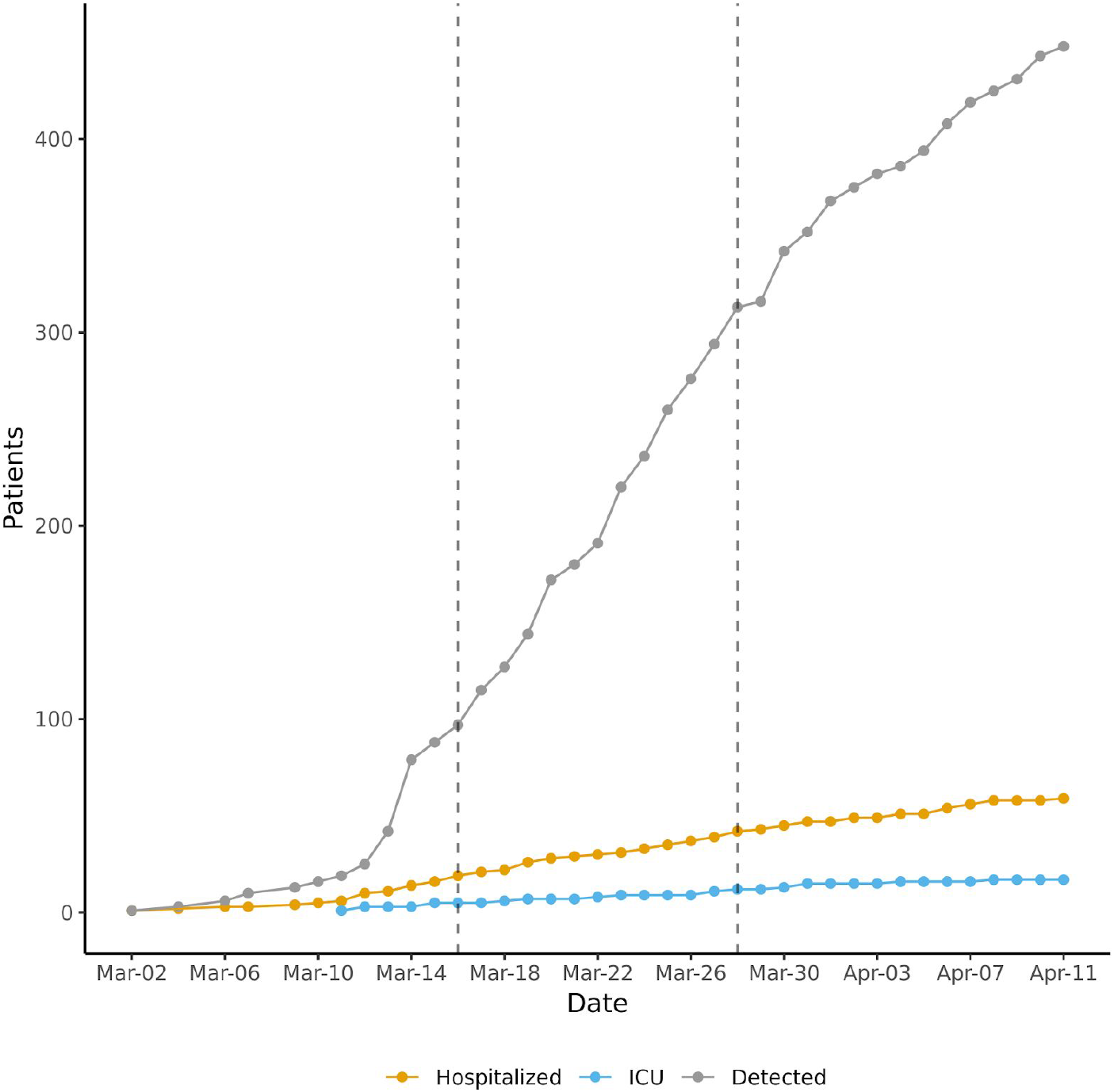
Slowed growth in total cases detected, hospitalized and admitted to ICU is seen after the SIP order (first dashed line). The prolongation of the doubling time of hospitalizations (yellow) happens faster and earlier than cases detected (gray). The divergence between the rate of cases detected and slower rate of hospitalizations is seen within 10 days of the SIP order. Doubling times calculated over a 7 day sliding window show that by March 28th (second dashed line), cases were doubling every 9 days but hospitalizations were doubling every 13 days.

Given that the test results and admissions data are available in nearly real time, health systems based monitoring of admission rates and the doubling time of hospitalized patient counts can provide accurate data for both public health planning and epidemiological modeling ^3^.

### Recently detected COVID-19 cases are younger than those detected prior to social distancing measures

As shown in Figure 3, between weeks 11 and 14 of the epidemic, there was no significant shift in the age distribution of patients tested. However, the average age of COVID-19 positive patients decreased (P-value = 0.0004) from 55.6 years [95% CI 53.0 - 58.3] prior to social distancing (March 16th, week 11) to 49.8 years [95% CI 47.9 - 51.7] after two weeks of social distancing (March 30th, week 14).

**Figure 3.**
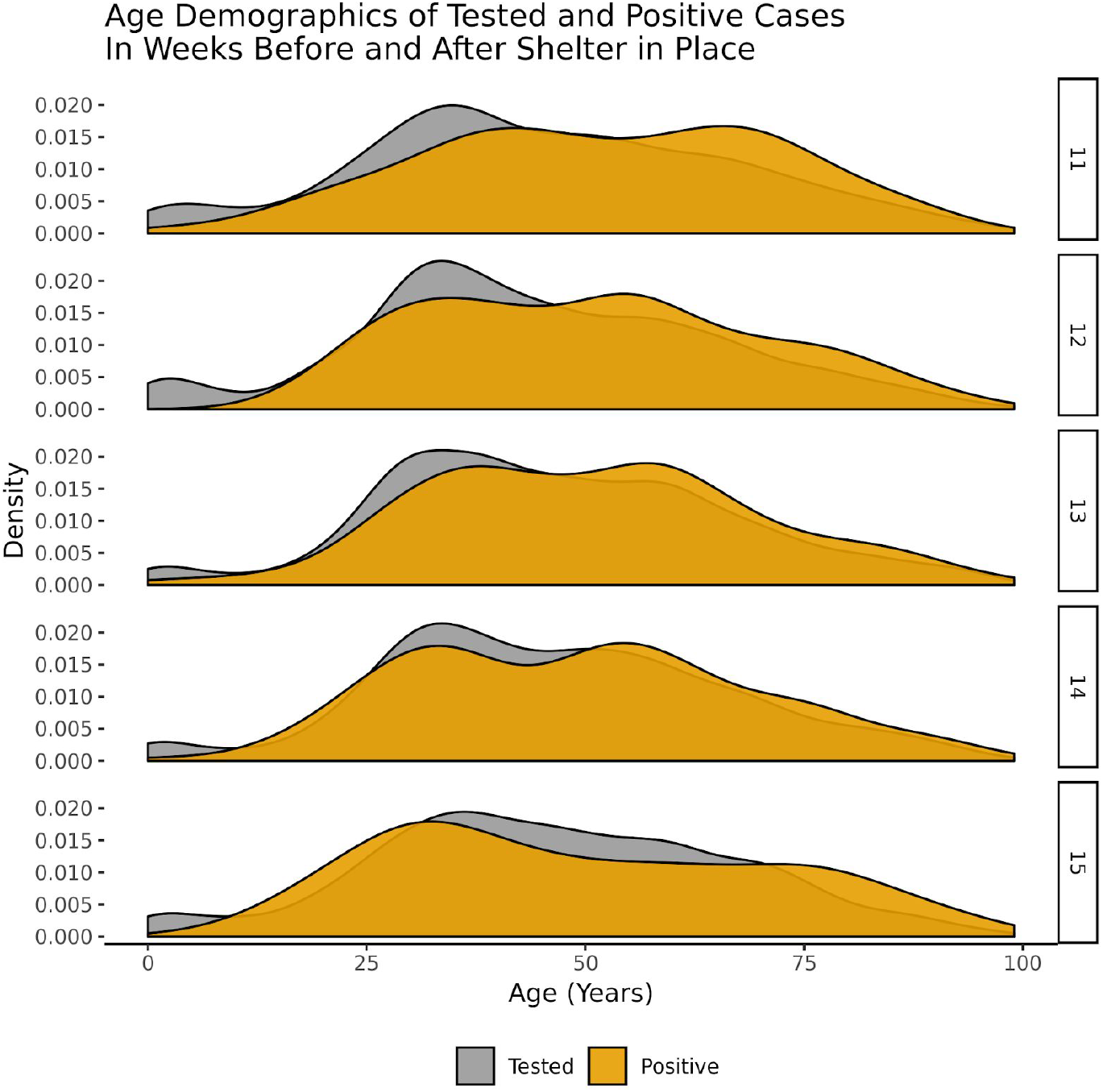
Change in age distribution of patients with positive SARS-CoV-2 test (brown) compared to those tested for SARS-CoV-2 (gray) for four weeks after sheltering in place order.

### Characteristics of COVID-19 cases hospitalized later are the same as those hospitalized earlier

Compared to before social distancing, the mean length of stay of hospitalized cases and rate of ICU admission was not significantly different (1.73 days shorter [95% CI −2.16, 5.62] and 4.1% absolute increase [95% CI −30%, 38%], respectively) than two weeks after social distancing (difference in means P-value = 0.378 and 0.810, respectively). Because the length of stay is right-censored for patients still in the hospital, these estimates are corrected for censoring.

**Table 1.** Characteristics of COVID-hospitalized cases around the state-mandated shelter-in-place (SIP) ordered on March 16th. Length of stay is adjusted for censoring using the Kaplan-Meier curve and 14 day restricted mean length of stay.

| Period | Hospitalized Cases | Age | Length of Stay Days | % Hospitalized Admitted to ICU | Transfer to ICU From Admission Days (sd) |
| --- | --- | --- | --- | --- | --- |
| Pre SIP | 16 | 58.1 (15.1) | 8.31 | 31.2 | 1.72 (1.4) |
| First two weeks SIP | 28 | 59 (18.7) | 6.91 | 25 | 1.19 (1.7) |
| >2 weeks SIP | 15 | 61.4 (19.1) | 6.58 | 33.3 | 0.76 (0.65) |

## Discussion

While most epidemic simulation models use new case rates, no currently published model takes into account the shifts in demographics of positive patients, which is a major determinant of future hospital admission rates. If most new cases are younger, the corresponding need for hospitalizations will also be lower. Over the course of five weeks, we did not find a significant change in the age distribution of patients presenting with influenza like illness (ILI) and tested for COVID-19, however we did find a significant shift towards younger patients in those testing positive for COVID-19. The simplest explanation for the shift towards younger patients testing positive could be relaxing testing criteria. For example, guidelines at our institution went from requiring ILI symptoms and a known COVID-19 exposure, to expressing symptoms consistent with ILI, to medical doctor discretion. However, this does not explain why there is no change in the demographics of patients getting *tested* for COVID-19. A more plausible explanation is that because of the SIP order, the at-risk elderly population is protected and hence less likely to contract COVID-19. This interpretation is supported by the fact that despite seeing a larger number of younger COVID-19 positive patients, those that need to be admitted have similar ages, rate of ICU admission, and length of stay as before the SIP order; now we see fewer absolute numbers of such cases.

Our analysis spanning 26 days after shelter-in-place clearly demonstrates that the rate of confirmed cases, hospitalizations and ICU admissions for COVID-19 has flattened. The decrease in the rate of new hospitalizations began within 10 days after shelter-in-place was initiated and continues today. Despite the decrease in hospitalizations, we continue to see new patients presenting with influenza like illness (ILI) and younger patients testing positive for COVID-19 indicating ongoing community spread; but perhaps in a lower risk population.

This analysis demonstrates how--compared to new case counts--new hospitalizations is a better metric both for detecting the effect of SIP and for estimating the anticipated burden on the health system ^10,11,18^. Our findings also suggest that existing surge planning efforts should frequently recompute hospitalization doubling time because the change can be swift as seen in our data^19^. Models that do not use local hospitalization rates as well as the age distribution of the positive patients are likely to overestimate the resource burden of COVID-19.

Investments made in setting up data feeds prior to and immediately at the outset of this crisis were critical to accessing such data quickly. Additionally, a unification of our IT organizations across the School of Medicine and the Healthcare System, which completed in September 2019, proved to be immensely valuable enabling the setup of data access (such as to the laboratory testing data) in a timely manner.

Given the relative ease of obtaining such data at a health system, these metrics can not only help quantify the effects of state-mandated SIP, but also enable better planning of health system capacity to aid any actions required to return to pre-crisis operations^20^. For any reopening scenario, accurate projections of near-term health system capacity requirements are essential^21^. Therefore, we must start using these easily available, and useful, inputs right away.

## Data Availability

Patient-level healthcare data are not publicly available. Code to reproduce technical aspects of the analysis is available from the corresponding author by request.

## Contributorship Statement

All authors were involved in drafting or critically revising the presented work, gave final approval of the submitted manuscript, and agree to be accountable for ensuring the integrity of the work. S.G., S.K., N.H.S. and S.Y. drafted the manuscript, which was reviewed and edited by all co-authors. S.G., S.K., N.H.S. conceived of the analysis design. B.A.P acquired, helped interpret and analyze lab testing data. J.A.F and A.C. analyzed hospitalized patients’ admission characteristics and presenting symptom severity. S.G, S.K and S.Y. conducted statistical analysis of the data, and S.Y. conducted additional statistical analyses to produce the censor-adjusted estimates.

## Acknowledgements

We acknowledge support from the Departments of Medicine and Pathology at Stanford.

